# Humoral response, associated symptoms and profile of patients infected by SARS-CoV-2 with taste or smell disorders in the SAPRIS multicohort study

**DOI:** 10.1101/2022.02.01.22270250

**Authors:** Julien Ramillon, Xavier de Lamballerie, Olivier Robineau, Helene Blanché, Gianluca Severi, Mathilde Touvier, Marie Zins, Fabrice Carrat, Nathanaël Lapidus, the SAPRIS-SERO study group

## Abstract

**Objectives:** Taste or smell disorders have been reported as strongly associated with COVID-19 diagnosis. We aimed to identify subject characteristics, symptom associations, and humoral response intensity associated with taste or smell disorders.

**Patients and methods:** We used data from SAPRIS, a study based on a consortium of five prospective cohorts gathering 279,478 participants in the French general population. In the analysis, we selected participants who were presumably infected by SARS-CoV-2 during the first epidemic wave.

**Results:** The analysis included 3,439 patients with a positive ELISA-Spike. Sex (OR = 1.28 [95% CI 1.05-1.58] for women), smoking (OR = 1.54 [95% CI 1.13-2.07]), consumption of more than 2 drinks of alcohol a day (OR = 1.37 [95% CI 1.06-1.76]) were associated with a higher probability of taste or smell disorders. The relationship between age and taste or smell disorders was non-linear. Serological titers were associated with taste or smell disorders: OR = 1.31 [95% CI 1.26-1.36], OR = 1.37 [95% CI 1.33-1.42] and OR = 1.34 [95% CI 1.29-1.39] for ELISA-Spike, ELISA-Nucleocapsid and seroneutralization, respectively. Among participants with taste or smell disorders, 90% reported a wide variety of other symptoms whereas 10% reported no other symptom or only rhinorrhea.

**Conclusion:** Among patients with a positive ELISA-Spike test, women, smokers and people drinking more than 2 drinks a day were more likely to develop taste or smell disorders. This symptom was strongly associated with a humoral response. The overwhelming majority of patients with taste or smell disorders experienced a wide variety of symptoms.

## 1. Introduction

Symptoms of COVID-19 are commonly cough, fever, dyspnea, myalgia, headache, arthralgia and diarrhea [1,2]. The spread of the epidemic highlighted a new disease presentation: patients with taste or smell disorders (TSD). This symptom has been reported to be highly predictive of COVID-19 diagnosis [3]. There is no consensus in the literature about patient characteristics associated with this symptom. There remain many unanswered questions about the relationships between immune response intensity and TSD and their association with other COVID-19 symptoms. Although long-term data in the general population are still scarce, it appears that TSD may be persistent over time in some patients [4].

To provide a better understanding of TSD in COVID-19 patients, we conducted an analysis based on data from the SAPRIS multicohort study. Our main goals were to identify subject characteristics associated with TSD, to investigate the relationship between TSD and immune response intensity and to characterize symptom associations for subjects with or without TSD.

## 2. Material and method

### 2.1. Patient and public involvement

This study was designed without patient or public involvement.

### 2.2. Design

We used data from SAPRIS (“Santé, Perception, pratiques Relations et Inégalités Sociales en population générale pendant la crise COVID-19”) based on a consortium of prospective cohort studies involving three general population-based adult cohorts: CONSTANCES, a “general population” cohort including a representative sample of 215,000 adults (including 66,000 followed by internet) aged 18 to 69 years at inclusion and recruited from 2012; E3N / E4N, a multigenerational adult cohort based on a community of families with 113,000 participants (women recruited in 1990 and still actively followed, their offspring and the fathers of this offspring) among whom 90,000 have been invited to an internet follow-up; and NutriNet-Santé a nutritional general population-based internet cohort started in 2009, with 170,000 included participants. Details on the SAPRIS survey are available elsewhere [5].

### 2.3. Participants and dates

All participants were invited to respond to two electronic self-administered questionnaires. Questionnaires were sent as of April 1, 2020 and returned before May 27, 2020, thus they covered the lockdown and post lockdown period (in France, the first lockdown occurred between March 17, 2020 and May 11, 2020). These data were combined with serological results from the SAPRIS-SERO study, in which a random sample of SAPRIS participants were invited to provide self-sampling dried-blood spot (DBS) aimed at serology testing [6].

Overall, 279,478 participants were invited to respond to SAPRIS questionnaires, 102,001 (37%) completed both questionnaires, and among them, 93,610 were invited to perform the serology, 86,913 (93%) returned dried blood spot and a serology could be performed and interpretable in 82,787.

In this analysis, we selected participants who had been presumably infected by SARS-CoV-2 based on serological results, i.e. all participants with a positive ELISA-S result, leading to a set of 3,695 participants.

Ethical approval and written or electronic informed consent were obtained from each participant before enrolment in the original cohort. The SAPRIS survey was approved by the Inserm ethics committee (approval #20-672 dated March 30, 2020). The SAPRIS-SERO study was approved by the Sud-Mediterranee III ethics committee (approval #20.04.22.74247) and electronic informed consent was obtained from all participants for DBS testing.

### 2.4. Data sources/measurement

Symptoms were reported if they had been present at least once within 14 days prior to each questionnaire. Smoking status, alcohol consumption and body mass index (BMI) were extracted from the original cohort databases and updated in 2020 with self-reported questionnaires.

The ELISA test (Euroimmun®, Lübeck, Germany) was used to detect anti-SARS-CoV-2 antibodies (IgG) directed against the S1 domain of the spike protein of the virus (ELISA-S). Following the manufacturer’s instructions, an ELISA-S test was considered to be positive with an optical density ratio ≥ 1.1, indeterminate between 0.8 and 1.1, and negative < 0.8. The sensitivity and specificity of the ELISA-S test at the 1.1 threshold (considering indeterminate results as negative) were reported to be 87% and 97.5%, respectively [7].

All samples with an ELISA-S test ≥ 0.7 were also tested with an ELISA test to detect IgG antibodies against the SARS-CoV-2 nucleocapsid protein (Euroimmun®, Lübeck, Germany, ELISA-NP) using the same thresholds as above and with an in-house micro-neutralization assay to detect neutralizing anti-SARS-CoV-2 antibodies (SN), as described elsewhere with a positive SN defined as a titer ≥ 40 (SN titer was determined by iterative dilutions, leading to discrete values for titers, equal to 10, 20, 40, 80 or 160) [8].

### 2.5. Variables

The main outcome was the presence of self-reported TSD on either the first or the second questionnaire. Systemic symptoms were defined as presence of fever or muscular aches or headaches; digestive symptoms as presence of diarrhea or nausea; pulmonary symptoms as presence of cough, dyspnea or chest pain. Smoking status was defined as a binary variable “active smoker” versus “Non-smoker or former smoker”. Alcohol consumption was defined as a binary variable “≤ 2 drinks per day” versus “> 2 drinks per day”. BMI was defined as a binary variable “overweighted or obese (BMI ≥ 25 kg/m^2^)” versus “underweighted or normal (BMI < 25 kg/m^2^)”.

We considered the optical density ratios of ELISA-S and ELISA-NP as well as neutralizing anti-SARS-CoV-2 antibody titers as quantitative variables. To compensate for the non-normality of the distribution, statistical analyses were conducted on log-transformed titers.

### 2.6. Missing data

Participants with missing data on the main outcome or other covariates were excluded from the analyses. A sensitivity analysis with multiple imputation of missing data was conducted to assess robustness of results: 20 imputed datasets were generated with multiple imputation by chained equations (predictive mean matching), among which all estimates were poled using Rubin’s rule.

### 2.7. Statistical method

Logistic regression models were used to identify factors associated with TSD. These models considered sex, age, smoking status, alcohol consumption and BMI. To account for nonlinearity, age was included in the regression models using restricted cubic splines. We also tested interactions between sex and either smoking status or alcohol consumption. Interaction terms and nonlinear terms for age were retained in the final model if they improved the BIC criterion.

Logistic regression models were used to estimate the association between TSD and serological titers according to the three assays, respectively. Analyses were adjusted for age, sex, smoking status, alcohol consumption and BMI, as potential cofounders, in each model. Associations with TSD were reported as odds ratios (OR) with their 95% confidence intervals (CI).

Marginal and joint distributions of symptoms were reported with upset plots. For the group of patients with TSD, the TSD symptom was included in the analysis whereas in the group without TSD, an “asymptomatic” category was created and included in the analysis. Both analyses included digestive, respiratory, systemic, fatigue and rhinorrhoea symptoms. Hierarchical ascending classification was used to identify subsamples of subjects with similar symptoms associations.

All analyses were conducted with the R statistical software version 4.0.3. Reporting of this research follows STROBE guidelines [9].

## 3. Results

Out of the 93,610 participants who completed both questionnaires and were invited to perform a DBS, 3,439 with a positive ELISA-S with available information on TSD were included in the analysis. A detailed flow diagram is provided (Figure 1). Sensitivity analyses relying on multiple imputation provided similar results whenever conducted (results not shown).

**Figure 1.**
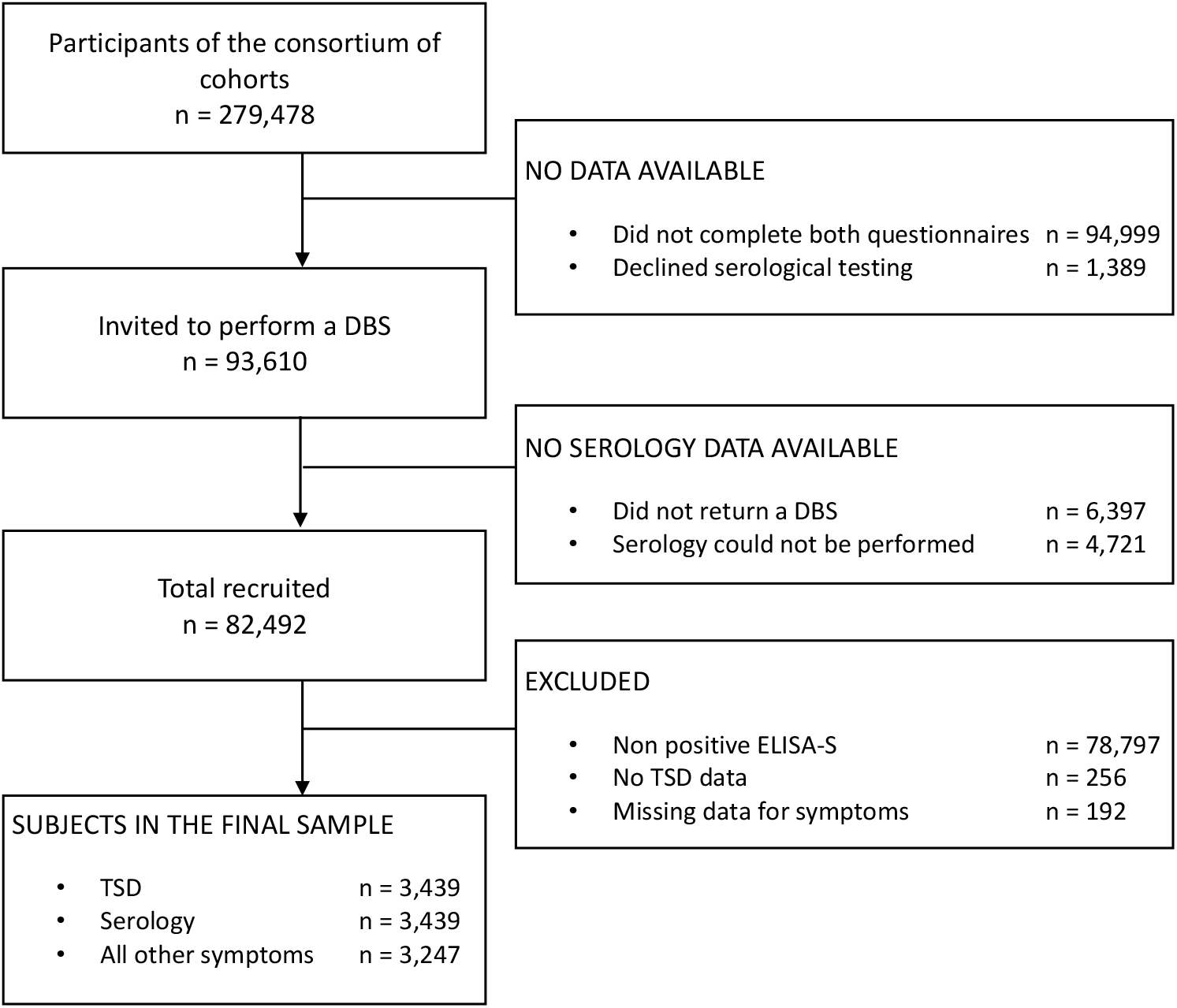
Inclusion of participants in the study. Flow diagram describing the process of inclusion in our study for participants to SAPRIS cohort.

### 3.1. Patients’ characteristics and association with TSD

Table 1 presents the characteristics of participants with positive ELISA-S (PE-S), along with ORs from univariable and multivariable analyses. Out of the 3,439 PE-S participants, 750 (21.8%) reported TSD. Proportions of women were 71.2% and 68.7% in participants with and without TSD, respectively. Median ages were 51 and 46 years.

**Table 1:** Characteristics of participants with and without taste or smell disorders (TSD). Categorial variables are reported as counts (percentages of non-missing values) and quantitative variables as median [Q1 – Q3]

|  | Characteristics of participants with positive ELISA-S |  | Univariable analysis |  | Multivariable analysis |  |
| --- | --- | --- | --- | --- | --- | --- |
|  | With TSD<br>(n = 750) | Without TSD<br>(n = 2689) | OR | 95% CI | OR | 95% CI |
| Sex |  |  |  |  |  |  |
| Men | 216 (28.8%) | 841 (31.3%) | 1 |  | 1 |  |
| Women | 534 (71.2%) | 1848 (68.7%) | 1.13 | 0.94 - 1.35 | 1.28 | 1.05 - 1.58 |
| Smoking status |  |  |  |  |  |  |
| Non-smoker or former smoker | 628 (87.7%) | 2416 (91.3%) | 1 |  | 1 |  |
| Active smoker | 88 (12.3%) | 229 (8.7%) | 1.48 | 1.13 - 1.91 | 1.54 | 1.13 - 2.07 |
| Missing data | 34 | 44 |  |  |  |  |
| Alcoholic consumption |  |  |  |  |  |  |
| 2 or less drinks a day | 554 (82.0%) | 2153 (86.6%) | 1 |  | 1 |  |
| 3 or more drinks a day | 122 (18.0%) | 333 (13.4%) | 1.42 | 1.13 - 1.78 | 1.37 | 1.06 - 1.76 |
| Missing data | 74 | 203 |  |  |  |  |
| BMI |  |  |  |  |  |  |
| Underweighted or normal (BMI < 25) | 457 (64.3%) | 1690 (65.0%) | 1 |  | 1 |  |
| Overweighted or obese (BMI ≥ 25) | 254 (35.7%) | 912 (35.0%) | 1.03 | 0.87 - 1.22 | 1.01 | 0.83 - 1.22 |
| Missing data | 39 | 87 |  |  |  |  |
| Age |  |  |  |  |  |  |
| Age | 51 [42 - 61] | 46 [40 - 60] |  |  |  |  |
| 30 years old |  |  | 1.97 | 1.45 – 2.69 | 1.82 | 1.27 – 2.60 |
| 40 years old |  |  | 1 |  | 1 |  |
| 50 years old |  |  | 1.77 | 1.54 – 2.04 | 1.91 | 1.63 – 2.23 |
| 60 years old |  |  | 3.70 | 3.07 – 4.45 | 4.33 | 3.54 – 5.29 |
| 70 years old |  |  | 1.58 | 1.29 – 1.92 | 1.67 | 1.35 – 2.05 |

Multivariable analyses included 3,439 participants with PE-S. No interaction was found between sex and either smoking status or alcohol consumption. Nonlinear terms for age improved the BIC criterion and were kept in the selected model. Sex (OR = 1.28 [95% CI 1.05-1.58] for women), smoking (OR = 1.54 [95% CI 1.13-2.07]), alcohol consumption of at least 2 drinks per day (OR = 1.37 [95% CI 1.06-1.76]) were associated with a higher probability of TSD. Compared with subjects aged 40 years as a reference, those aged 30 (OR = 1.82 [95% CI 1.27-2.60]), 50 (OR = 1.91 [95% CI 1.63-2.23]), 60 (OR = 4.33 [95% CI 3.54-5.29]) or 70 (OR = 1.67, [95% CI 1.35-2.05]) had a higher probability of reporting TSD. The nonlinear age-dependent estimated OR is reported in Figure 2.

**Figure 2.**
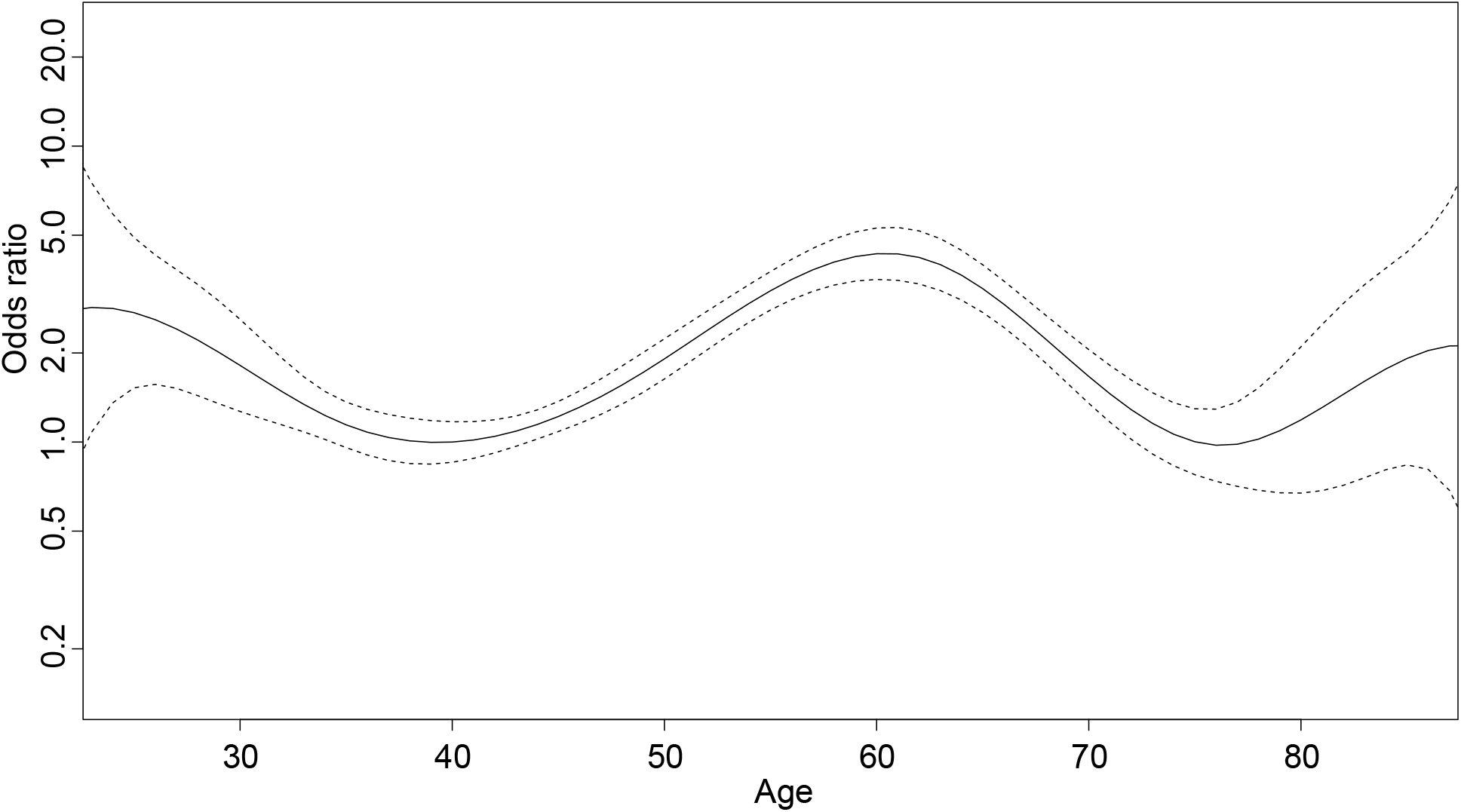
Age-dependent estimated OR (and 95% CI) of taste or smell disorder, people aged 40 as reference. Graphic representation of age-dependent estimated OR (and 95% CI) of taste and smell disorders in multivariable analyses. People aged 40 are considered as reference.

### 3.2. Serology

Distributions of serological titers in PE-S participants are reported in Table 2. Associations between serological titers and TSD were estimated in three independent multivariable models. Related ORs are reported per 0.1 augmentation in the log-transformed titers and can approximately be interpreted as ORs per 10% increase in serological titers. All titers were associated with a higher probability of TSD: OR = 1.31 [95% CI 1.26-1.36], OR=1.37 [95% CI 1.33-1.42] and OR=1.34 [95% CI 1.29-1.39] for ELISA-S, ELISA-NP and seroneutralization, respectively.

**Table 2:** Distribution of serological titers and associations with taste or smell disorders (TSD) adjusted for sex, age, smoking status, alcohol consumption and BMI

|  | With TSD<br>(n = 750) |  | Without TSD<br>(n = 2689) |  | Multivariable<br>analysis |  |
| --- | --- | --- | --- | --- | --- | --- |
|  | Median | Q1 - Q3 | Median | Q1 - Q3 | OR | 95% CI |
| ELISA-S | 1.90 | 1.40 - 2.97 | 2.95 | 1.88 - 4.79 | 1.31 | 1.26 - 1.36 |
| ELISA-NP | 0.60 | 0.38 - 1.37 | 2.19 | 1.28 - 3.79 | 1.37 | 1.33 - 1.42 |
| SN | 10 | 10 - 20 | 40 | 20 - 160 | 1.34 | 1.29 - 1.39 |

### 3.3. Symptoms

Analyses regarding the association of TSD with other symptoms included 3,247 participants with PE-S and no missing data for symptoms. Table 3 reports results of the hierarchical clustering analysis, using only reported symptoms in PE-S participants. The number of clusters was arbitrarily set to 4. Cluster #1 groups all asymptomatic participants; cluster #2 groups participants with various associations of symptoms; cluster #3 groups participants with TSD, mostly isolated or associated with rhinorrhea; whereas cluster #4 groups patients with digestive symptoms, mostly isolated or associated with fatigue. Table 3 also presents the distribution of age, sex and serological titers in these clusters. These characteristics were not used to identify clusters.

**Table 3:**
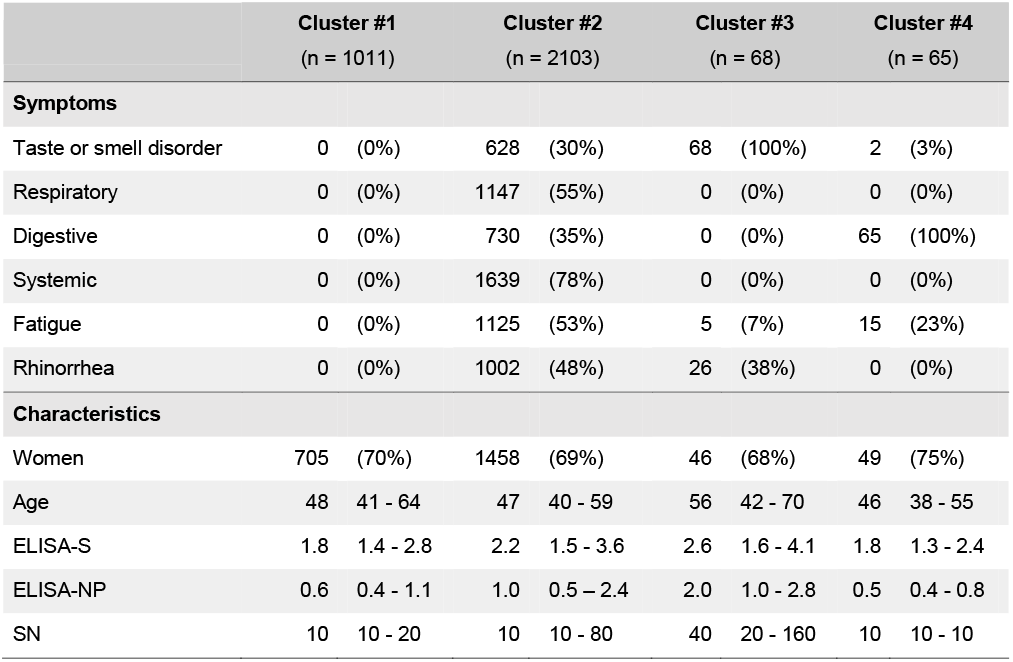
Characteristics of participants depending on symptoms profile. Categorical variables are reported as counts (percentages) and quantitative variables as median [Q1 – Q3]

|  | Cluster #1 |  | Cluster #2 |  | Cluster #3 |  | Cluster #4 |  |
| --- | --- | --- | --- | --- | --- | --- | --- | --- |
|  | (n = 1011) |  | (n = 2103) |  | (n = 68) |  | (n = 65) |  |
| Symptoms |  |  |  |  |  |  |  |  |
| Taste or smell disorder | 0 | (0%) | 628 | (30%) | 68 | (100%) | 2 | (3%) |
| Respiratory | 0 | (0%) | 1147 | (55%) | 0 | (0%) | 0 | (0%) |
| Digestive | 0 | (0%) | 730 | (35%) | 0 | (0%) | 65 | (100%) |
| Systemic | 0 | (0%) | 1639 | (78%) | 0 | (0%) | 0 | (0%) |
| Fatigue | 0 | (0%) | 1125 | (53%) | 5 | (7%) | 15 | (23%) |
| Rhinorrhea | 0 | (0%) | 1002 | (48%) | 26 | (38%) | 0 | (0%) |
| Characteristics |  |  |  |  |  |  |  |  |
| Women | 705 | (70%) | 1458 | (69%) | 46 | (68%) | 49 | (75%) |
| Age | 48 | 41 - 64 | 47 | 40 - 59 | 56 | 42 - 70 | 46 | 38 - 55 |
| ELISA-S | 1.8 | 1.4 - 2.8 | 2.2 | 1.5 - 3.6 | 2.6 | 1.6 - 4.1 | 1.8 | 1.3 - 2.4 |
| ELISA-NP | 0.6 | 0.4 - 1.1 | 1.0 | 0.5 – 2.4 | 2.0 | 1.0 - 2.8 | 0.5 | 0.4 - 0.8 |
| SN | 10 | 10 - 20 | 10 | 10 - 80 | 40 | 20 - 160 | 10 | 10 - 10 |

Results are also presented using two upset plots (Supplementary figure S1): one for participants with TSD (Fig. S1A; n = 698) and one for participants without TSD (Fig. S1B; n = 2,549). Both plots took into account self-reported symptoms in at least one of the two questionnaires.

## 4. Discussion

Taste or smell disorders have been extensively reported as common symptoms in SARS-CoV-2 infections [10]. Direct contact and interaction of the virus with gustatory or olfactory receptor cells may be the main cause for this symptom, though other pathophysiologic pathways are possible [11].

### 4.1. Key findings

We found that the risk of reporting TSD for participants with a positive SARS-CoV-2 ELISA-S test is higher in women, smokers and participants drinking more than 2 drinks of alcohol a day, after adjustment for age. The association with age was nonlinear, with a lower probability of TSD for participants aged around 40, compared with younger or older ones. In addition, our analyses showed a strong association between developing TSD and an intense immune response for people with a positive ELISA-S. Regarding symptoms, our study shows two patterns of symptom association in participants with TSD: in 90% of them, TSD are associated with a wide variety of symptoms, while in 10% of them, TSD are isolated or associated with rhinorrhea.

### 4.2. Interpretation

#### 4.2.1. Patient profile

The association between TSD and female gender in COVID-19 infected subjects is well documented [12–14] and previous findings support our results. The nonlinear association with age, suggesting that patients around 40 years present a lower probability to report TSD, is not easily interpretable as conflicting results were reported regarding the association of TSD with age: a study showed that age higher than 18 years was associated with TSD, compared to 15-17 years, with results suggesting a higher frequency of TSD in infected subjects aged 18-44 years than adolescents and older subjects [12]. Another study found a lower prevalence of TSD in elderly patients [15]. Regarding the association we found between smoking status and TSD, previous studies also reported conflicting results, some of them finding a similar association [16,17] while others did not [15,18,19]). A possible explanation for this association is that functional interactions between nicotine and the angiotensin-converting enzyme 2 (ACE2) facilitate the infection of cells by the virus [20]. To our knowledge, the association between TSD and alcoholic consumption was reported – and nonsignificant – in only one case-control study with a limited sample size [21]. No obvious biological mechanism is likely to explain this result and a spurious association due to residual confounding cannot be ruled out. The non-association between TSD and overweight is consistent with the literature [15].

#### 4.2.2. Immune response

We found that the development of TSD was strongly associated with the humoral response. Even though the pathophysiology of TSD in COVID-19 infection is still poorly understood, these results, combined with the literature, suggest the existence of a biological mechanism between immune response and TSD. Animal studies suggest that the interaction between ACE2 and the spike protein may yield to massive infection of sustentacular cells in the olfactory epithelium and immune cell infiltration leading to global desquamation of this epithelium [22,23]. Other studies support that cell infection by the SARS-CoV-2 and infiltration of immune cells in the olfactory epithelium could lead to olfactory sensory neurons infection by horizontal spreading [24] or to loss of odorant receptor [25].

#### 4.2.3. Symptoms

We discerned different patterns of symptoms associations in participants with a positive ELISA-S test. Participants with TSD were more likely to report a wide variety of symptoms, while most participants without TSD reported a complete absence of symptoms or isolated symptoms (systemic or rhinorrhea mainly). In the literature, the majority of patients with TSD experienced at least one other symptom [15,26,27], which is consistent with our findings.

### 4.3. Strengths and limitations

Our study presents numerous strengths. Participants from the SAPRIS study were recruited from the general population, via well-characterized cohorts with a very high participation rate. This design ensured a large sample size and a more comprehensive overview on COVID-19 symptoms than studies focusing only on either outpatients or inpatients. Moreover, our case definition is prospective and does not rely on reported symptoms, hence the important number of asymptomatic or paucisymptomatic patients.

Regarding serology, most DBS were collected within 1 to 3 months following a period of intense viral circulation and serological analyses were centralized and performed blinded to subjects and investigators. The combination of several serological analysis methods (ELISA-S, ELISA-NP and seroneutralization) tends to reinforce the robustness of the results regarding the association between humoral response and TSD.

Several limitations must also be noted. First, we considered subjects with a positive ELISA-S (i.e. ≥ 1.1) as having been infected by the SARS-CoV-2. This choice may tend to underestimate the number of subjects who have been infected with the COVID 19 virus as (i) indeterminate results (between 0.8 and 1.1) were considered as negative, (ii) there is evidence that the humoral immunity developed in the weeks following infection decreases over time (though this issue was partly controlled as DBS were mostly collected soon after the first epidemic wave) [28] and (iii) 10% to 20% of infected individuals will not mount a detectable humoral response [29,30]. Thus, though this case definition favoured specificity over sensitivity and limits the occurrence of false positive cases, we may have selected infected participants whose initial humoral response was intense while infected participants with a milder immune response may have not been detected. As only one sample was analysed per participant, within-participant dynamics of the serological response could not be studied. Second, TSD, as well as all other symptoms, were self-reported. It has been shown that self-reporting of TSD may lead to under-reporting of this symptom, compared with objective assessment by a professional [26]. Moreover, we may think that participants who reported a TSD are those who are the most likely to report other types of symptoms, which may partly explain the frequent associations of TSD with several other symptoms.

## 5. Conclusion

In conclusion, our study shows that among patients with a positive serology ELISA-S test, women, smokers and people drinking more than 2 drinks a day were more likely to develop TSD. We also found a strong association between humoral response and TSD. Regarding symptoms associations, the overwhelming majority of participants with TSD experienced a wide variety of acute symptoms, while a few of them only developed TSD either isolated or associated with only rhinorrhea.

## Data Availability

The datasets used and analyzed during the current study are available from the corresponding author on reasonable request.

## Acknowledgments

The authors warmly thank all the volunteers of the Constances, E3N-E4N, and NutriNet-Sante cohorts.

We thank the staff of the Constances, E3N-E4N and NutriNet-Sante cohorts that have worked with dedication and engagement to collect and manage the data used for this study and to ensure continuing communication with the cohort participants. We thank the CEPH-Biobank staff for their adaptability and the quality of their work. In the virology department, Dr Nadege Brisbarre and the technical staff for impeccable management of samples and serological assays.

## Ethical Approval

All procedures performed in studies involving human participants were in accordance with the 1964 Helsinki declaration and its later amendments.

## Funding

### This study

ANR (Agence Nationale de la Recherche, #ANR-20-COVI-000, #ANR-10-COHO-06), Fondation pour la Recherche Medicale (#20RR052-00), Inserm (Institut National de la Sante et de la Recherche Medicale, #C20-26).

### Cohorts funding

The CONSTANCES Cohort Study is supported by the Caisse Nationale d’Assurance Maladie (CNAM), the French Ministry of Health, the Ministry of Research, the Institut national de la sante et de la recherche medicale. CONSTANCES benefits from a grant from the French National Research Agency [grant number ANR-11-INBS-0002] and is also partly funded by MSD, AstraZeneca, Lundbeck and L’Oreal. The E3N-E4N cohort is supported by the following institutions: Ministere de l’Enseignement Superieur, de la Recherche et de l’Innovation, INSERM, University Paris-Saclay, Gustave Roussy, the MGEN, and the French League Against Cancer. The NutriNet-Sante study is supported by the following public institutions: Ministere de la Sante, Sante Publique France, Institut National de la Sante et de la Recherche Medicale (INSERM), Institut National de la Recherche Agronomique (INRAE), Conservatoire National des Arts et Metiers (CNAM) and Sorbonne Paris Nord University. The CEPH-Biobank is supported by the ≪ Ministere de l’Enseignement Superieur, de la Recherche et de l’Innovation ≫.

## Appendix

**Figure A.1.**
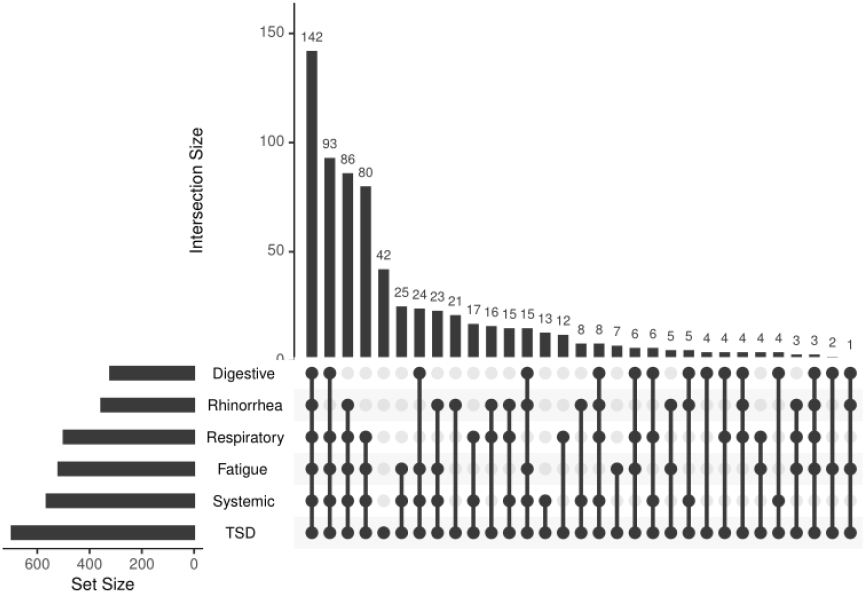
Symptoms associations in participants with taste or smell disorders. Graphic representation of symptoms associations by means of upset plot for participants with a positive ELISA-S and taste or smell disorders.

**Figure A.2.**
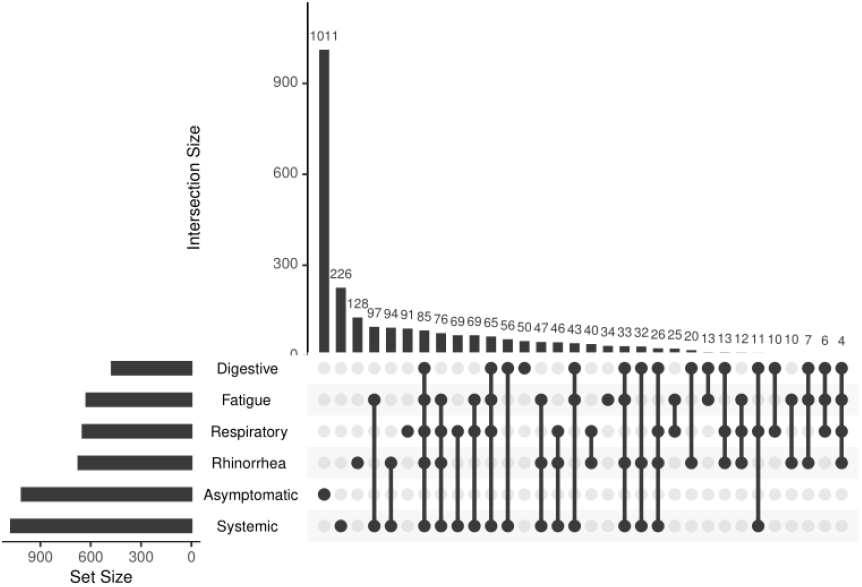
Symptoms associations in participants without taste or smell disorders. Graphic representation of symptoms associations by means of upset plot for participants without a positive ELISA-S and taste or smell disorders.

## Author statement

JR: Design, methodology, writing-all stage

XL: Conceptualisation, design, data gathering, writing-reviewing and editing

OR, HB, GS, MT, MZ: Data gathering, writing-reviewing and editing

FC: Conceptualisation, design, methodology, data gathering, writing-reviewing and editing

NL: FC: Conceptualisation, design, methodology, data gathering, writing-reviewing and editing

## The SAPRIS study group

Nathalie Bajos (co-Principal investigator), Fabrice Carrat (co-Principal investigator),Pierre-Yves Ancel, Marie-Aline Charles, Florence Jusot, Claude Martin, Laurence Meyer, Ariane Pailhe, Gianluca Severi, Alexis Spire, Mathilde Touvier, Marie Zins.

## The SAPRIS-SERO study group

Fabrice Carrat (Principal investigator), Pierre-Yves Ancel, Marie-Aline Charles, Gianluca Severi, Mathilde Touvier, Marie Zins

Sofiane Kab, Adeline Renuy, Stephane Le-Got, Celine Ribet, Emmanuel Wiernik,Marcel Goldberg, Marie Zins (Constances cohort),

Fanny Artaud, Pascale Gerbouin-Rerolle, Melody Enguix, Camille Laplanche, Roselyn Gomes-Rima, Lyan Hoang, Emmanuelle Correia, Alpha Amadou Barry, Nadege Senina, Gianluca Severi (E3N-E4N cohort)

Fabien Szabo de Edelenyi, Nathalie Druesne-Pecollo, Younes Esseddik, Serge Hercberg, Mathilde Touvier (NutriNet-Sante cohort)

Marie-Aline Charles, Pierre-Yves Ancel, Valerie Benhammou, Anass Ritmi, Laetitia Marchand, Cecile Zaros, Elodie Lordmi, Adriana Candea, Sophie de Visme, Thierry Simeon, Xavier Thierry, Bertrand Geay, Marie-Noelle Dufourg, Karen Milcent (Epipage2 and Elfe child cohorts)

Clovis Lusivika-Nzinga, Gregory Pannetier, Nathanael Lapidus, Isabelle Goderel, Celine Dorival, Jerome Nicol, Fabrice Carrat (IPLESP – methodology and coordinating data center)

Cindy Lai, Helene Esperou, Sandrine Couffin-Cadiergues (Inserm) Jean-Marie Gagliolo (Institut de Sante Publique)

Helene Blanché, Jean-Marc Sebaoun, Jean-Christophe Beaudoin, Laetitia Gressin, Valerie Morel, Ouissam Ouili, Jean-Francois Deleuze (CEPH-Biobank)

Stephane Priet, Paola Mariela Saba Villarroel, Toscane Fourie, Souand Mohamed Ali, Abdenour Amroun, Morgan Seston, Nazli Ayhan, Boris Pastorino, Xavier de Lamballerie (Unite des Virus Emergents)

